# The projected impact of mitigation and suppression strategies on the COVID-19 epidemic in Senegal: A modelling study

**DOI:** 10.1101/2020.07.03.20144949

**Authors:** Hayley A Thompson, Aminata Mboup, Badara Cisse, Shevanthi Nayagam, Oliver J Watson, Charles Whittaker, Patrick G T Walker, Azra C Ghani, Souleymane Mboup, With the Imperial College COVID-19 Response Team

## Abstract

**Background:** Physical distancing measures that reduce social contacts have formed a key part of national COVID-19 containment and mitigation strategies. Many Sub-Saharan African nations are now facing increasing numbers of cases of COVID-19 and there is a need to understand what levels of measures may be required to successfully reduce transmission.

**Methods:** We collated epidemiological data along with information on key COVID-19 specific response policies and health system capacity estimates for services needed to treat COVID-19 patients in Senegal. We calibrated an age-structured SEIR model to these data to capture transmission dynamics accounting for demography, contact patterns, hospital capacity and disease severity. We simulated the impact of mitigation and suppression strategies focussed on reducing social contact rates.

**Results:** Senegal acted promptly to contain the spread of SARS-CoV-2 and as a result has reduced the reproduction number from 1.9 (95% CI 1.7-2.2) to 1.3 (95% CI 1.2-1.5), which has slowed but not fully interrupted transmission. We estimate that continued spread is likely to peak in October, and to overwhelm the healthcare system with an estimated 77,400 deaths (95% CI 55,270-100,700). Further reductions in contact rates to suppress transmission (R_t_<1) could significantly reduce this burden on healthcare services and improve overall health outcomes.

**Conclusions:** Our results demonstrate that Senegal has already significantly reduced transmission. Enhanced physical distancing measures and rapid scale up of hospital capacity is likely to be needed to reduce mortality and protect healthcare infrastructure from high levels of demand.

## Background

COVID-19 disease, caused by the SARS-CoV-2 virus, represents an unprecedented public health emergency. Since the emergence of the virus in China at the end of 2019, worldwide spread has been rapid, with 7,823,289 cases and 431,541 deaths reported as of 15^th^ June 2020.^1^ The growing burden of COVID-19 in Sub Saharan Africa (SSA) is of particular concern given the relative lack of availability of healthcare. Experience to date has highlighted the substantial pressure COVID-19 epidemics place on health systems, leading to huge demand for therapeutic oxygen and mechanical ventilators, sufficient to overwhelm even comparatively well-resourced settings such as the US where there are an estimated 20-31 intensive care unit (ICU) beds per 100,000 people. In contrast, recent estimates suggest less than 1 ICU bed and 1 ventilator per 100,000 people across SSA.^2^

In Senegal, following the identification of an imported case, the government set up a response plan covering the period February to July 2020 with several modular phases depending on the epidemiological situation at the time. As of 15^th^ June, Senegal had recorded 5,173 confirmed cases, making it the fourth worst affected country in the Western African region.^3^ Prevention and control measures taken have included closure of the international airport (20^th^ March) and the declaration of a state of emergency (23^rd^ March) which included the closure of schools and mosques, banning of large public gatherings, travel restrictions and strengthening control over borders.^4^ In addition, funding for a multisectoral surveillance action/response plan was established. This plan aims to encourage people to adopt the right behaviours through communication strategies but also to strengthen the health system and initiate processes for monitoring activities. Initial testing was slow to scale-up, but now includes testing at health-centres and in the wider community in identified geographical hotspots, alongside contact tracing with isolation of confirmed symptomatic and asymptomatic cases, a total of 62,763 tests have been conducted since early March.^3^

Previous modelling work has highlighted that despite the younger populations in SSA, a combination of lower quality healthcare and large intergenerational family structures could result in significant morbidity and mortality unless the epidemic is swiftly brought under control.^5,6^ Whilst generic insights can be obtained from such modelling, country-specific models can be helpful during the early phase of the epidemic in order to guide changes in public health policy. Studies that have examined the potential epidemic trajectory in Senegal specifically are limited, with only two studies published that consider the current emergency measures and their impact on the transmission of the virus.^7,8^ Both of these studies estimate a significant final epidemic size with between 25-50% of the population infected, however neither of these studies address the impact that health systems capacity can have on COVID-19 related mortality.^5,7^ Here we describe the current epidemiological situation using data collected from across the country. We calibrate a dynamical model of COVID-19 transmission to these data to investigate the potential impact of different mitigation and suppression strategies on mortality and health system capacity and explore possible future trajectories of the still nascent epidemic in Senegal.

## Methods

### Epidemiological Data and Statistical Analysis

We use daily COVID-19 data collected by the Ministry of Health.^3^ To estimate the growth rate of the epidemic, we fit a log-linear model to daily reported case numbers from the period of early expontential growth (17^th^ April to 16^th^ May). The growth rate from the early phase of an outbreak can be linked to the initial reproduction number.^9^ We used the R package earlyR to estimate the reproduction number during the early phase of the outbreak in Senegal, noting we are using the date of a reported case as symptom onset data is not available.^10,11^

### Mathematical Model

We use a stochastic age-structured SEIR model (Additional file 1: Figure S1) described in Walker *et al*, available in the form of an R package.^6^ The model explicitly captures passage through different healthcare settings and includes both age-specific patterns of mixing (derived from contact surveys carried out in SSA and disease severity.^12,13^ In brief, upon infection, individuals enter a period of latent infection, after which they either develop mild infection (symptomatic or asymptomatic) or severe disease (pneumonia or acute respiratory distress) that requires hospitalisation. Severe cases either require treatment with high-pressure oxygen (captured here as hospital beds) or develop more severe disease that requires intensive care treatment and mechanical ventilation (captured here as ICU beds). The probabilities of an infected individual developing mild infection or severe disease are age dependent and based on estimates of the age-specific severity of COVID-19 from China and the UK.^13^ All those who recover from infection are assumed to be immune to re-infection for the duration of the epidemic. Within the model, hospital and ICU bed occupancy is dynamically tracked, with infected individuals requiring care assigned to either receive or not receive a bed based on those available at the given timepoint. Not receiving the appropriate care due to healthcare capacity being exceeded is assumed to increase the risk of mortality (Additional file 1: Figure S2, Table S1). To capture the impacts of a potentially weaker healthcare system on treatment outcomes we model the probability of dying across all age groups in non-severe cases that require and receive hospital treatment with high pressure oxygen as 20-30%, and 60% if this oxygen support is not available (Additional file 1: Table S1). See Walker *et al* for full model description.^6^ To parameterise capacity constraints, we use local estimates of healthcare capacity – of which there are 407 general hospital beds with oxygen supply and 35 ICU beds with mechanical ventilation available in the 5 major hospitals in Senegal.^14^

### Model Fitting

The model was calibrated to the time series of reported deaths in Senegal. We assume reported deaths provide the best indication of the stage of the epidemic. However, given that there is likely to be undocumented deaths in the community, we account for this by assuming a 50% death ascertainment rate. At the time of calibration (15^th^ June) reported deaths totalled 64. The stochastic model is fit to the daily deaths using a particle filter grid search and by allowing two parameters to vary; the start date of the epidemic and the initial R_0_ in the absence of interventions (See Additional file 1: Supplementary methods, for full description).

Given the emergency measures that were enacted on 23^rd^ March across Senegal, Google surveys show that there have been significant reductions in movement.^15^ By equally weighting the relative contributions of work and social changes relative to movements in early February we estimate an average 33% reduction in contacts as a result of the state of emergency (Additional file 1: Figure S3A). To reflect the changes movement patters will have on social contacts, we introduce a time varying reduction in the baseline R_0_. Given the limited evidence for increases in mobility data to associate proportionally with increases in transmission we assume that R_t_ does not change in response to the recent increases in movement patterns we have observed in the Google data.^16,17^ A sensitivity analysis is presented in Additional file 1: Figure S3B where we highlight the poorer model fit resulting if we assume proportional increases between movement and R_t_ after the 5^th^ May.

All analysis was performed using R version 3.6.3.^18^

### Model Projections

Following model fitting we examine a range of different intervention scenarios. In all forward projections, we implement new interventions from the date of calibration and run simulations for 2 years. We initially consider 4 scenarios that incorporate a range of assumptions regarding the degree of reduction in contacts (33% as our current estimate of the average effect of emergency measures and an enhanced reduction to 60%) and the period over which these reductions are sustained (3 months or indefinitely) (Table 1). Note that the 60% reduction in contacts is expected to translate to R_t_<1 given our estimates of R_0_ for Senegal (see Results) whereas the 33% reduction would keep R_t_>1.

**Table 1.**
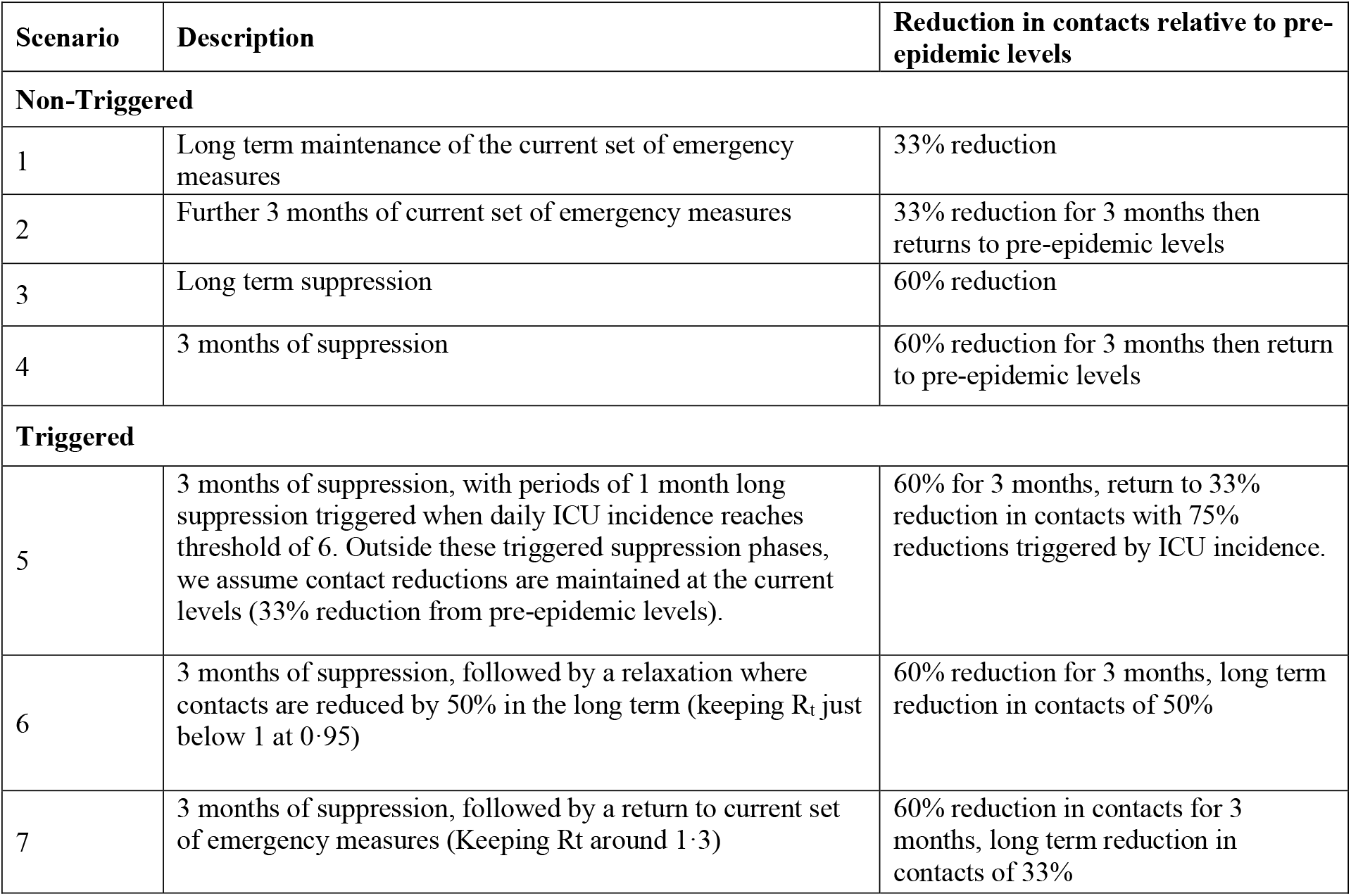
Description of mitigation and suppression strategies implemented across model projections.

| Scenario | Description | Reduction in contacts relative to pre-epidemic levels |
| --- | --- | --- |
| **Non-Triggered** |  |  |
| 1 | Long term maintenance of the current set of emergency measures | 33% reduction |
| 2 | Further 3 months of current set of emergency measures | 33% reduction for 3 months then returns to pre-epidemic levels |
| 3 | Long term suppression | 60% reduction |
| 4 | 3 months of suppression | 60% reduction for 3 months then return to pre-epidemic levels |
| **Triggered** |  |  |
| 5 | 3 months of suppression, with periods of 1 month long suppression triggered when daily ICU incidence reaches threshold of 6. Outside these triggered suppression phases, we assume contact reductions are maintained at the current levels (33% reduction from pre-epidemic levels). | 60% for 3 months, return to 33% reduction in contacts with 75% reductions triggered by ICU incidence. |
| 6 | 3 months of suppression, followed by a relaxation where contacts are reduced by 50% in the long term (keeping $R_t$ just below 1 at 0.95) | 60% reduction for 3 months, long term reduction in contacts of 50% |
| 7 | 3 months of suppression, followed by a return to current set of emergency measures (Keeping $R_t$ around 1.3) | 60% reduction in contacts for 3 months, long term reduction in contacts of 33% |

Given that longer-term suppression strategies are challenging to maintain, we also considered 3 further scenarios in which an initial period of suppression is followed by short periods of reduced contact rates to avoid health systems becoming overwhelmed (Table 1). We examine the impact of these interventions on estimated infections, deaths, and demand for hospital services.

## Results

### Current Epidemiological Situation

The first case of COVID-19 was reported in Senegal on 2^nd^ March, on 2^nd^ April, the first death was reported. As of 15^th^ June, there have been a total of 5,173 cases (Figure 1A) and 64 deaths, the majority of whom reported other underlying health conditions. Over time the distribution of cases reported has changed with the majority of cases classified as locally acquired by the 15^th^ June with 4,473 (86%) cases detected through contact tracing, 574 (12%) identified with no known epidemiological link and 126 (2%) cases reported as importations, with the majority of these cases (2894, 56%) reported in Dakar (Figure 1B). Similar proportions of cases have been reported in males (52%) and females (48%) and the highest proportion of cases are reported from those aged 20-39 years old (Figure 1C). 22 cases were being treated in critical care with most reported cases either exhibiting mild symptoms or asymptomatic, likely reflecting the primary detection route via contact tracing. This case distribution may also partly explain the relatively young age distribution of cases presenting in Senegal thus far.^3^

**Figure 1.**
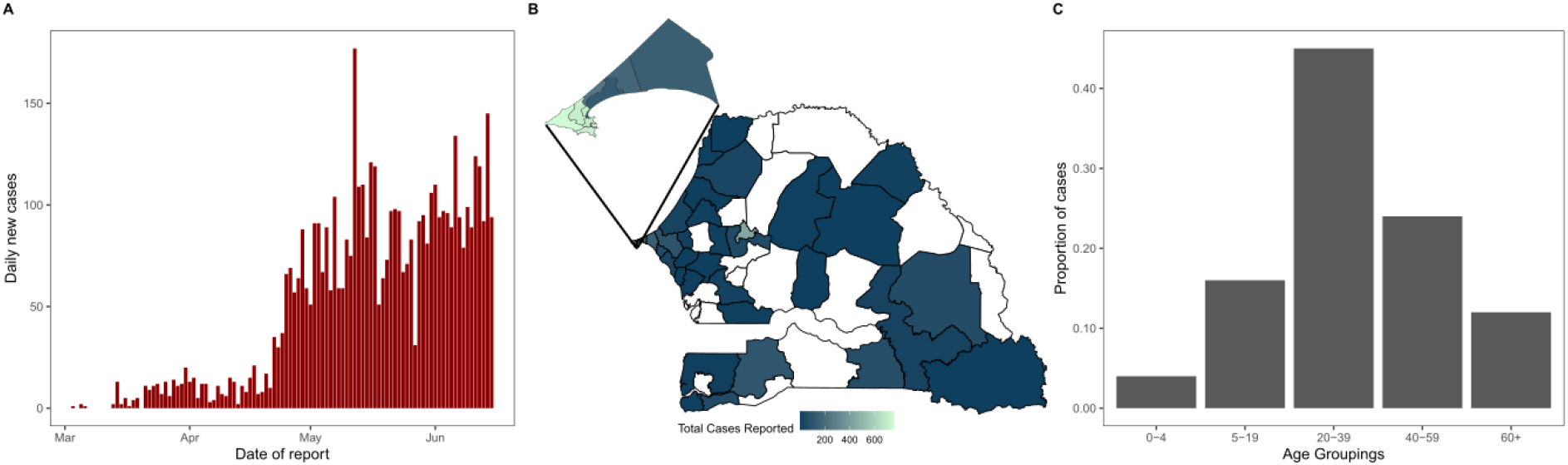
Epidemiology of COVID-19 in Senegal. **A)** Daily reported cases numbers for Senegal from the Ministry of Health up to 15^th^ June.^3^ **B)** The geographic location of cases in Senegal reported at the subnational department level as reported from the Ministry of health up to 15th June.^3^ **C)** Age distribution of cases last reported on the 4^th^ June.^38^

Using reported cases, we estimate a doubling time of 7·7 days (95% CI 6·2–9·9) based on a fitted growth rate of 0·09 per day (95% CI 0·06–0·11). Under an assumed serial interval of 6·5 days this translates to an estimate of R_t_ of 1·3 (95% CI 1·2–1·5).^19^ Such estimates are however, highly sensitive to the testing policy and therefore this may be biased. Furthermore, this estimate represents an average reproduction number over the early period of the epidemic, which will incorporate both the underlying R_0_ and the impact of early interventions.

Following model fitting our best estimate of the start date of the epidemic is 18^th^ February with a baseline R_0_ estimate of 1·9 (95% CI 1·7–2·2) (Figure 2A), which following the introduction of the emergency measures reduced to Rt=1·3 (95% CI 1·2–1·5). This estimate is in line with the estimate from the raw case data which only captures growth after the introduction of measures. Based on the 64 reported deaths used to calibrate the model we estimate that 37,603 (95% CI 26,565–53,571) people will have been infected with SARS-CoV-2 between 25^th^ February and 15^th^ June. It is important to note that approximately 50% of these infections are likely to have been asymptomatic.^13^ With 5,173 cases reported up to 15^th^ June, we estimate a national infection ascertainment fraction of 13·8% (95% CI 9·8%-19.4%). Importantly there is considerable uncertainty around our estimates and all future projections, given the small number of deaths reported to date Figure 2.

**Figure 2.**
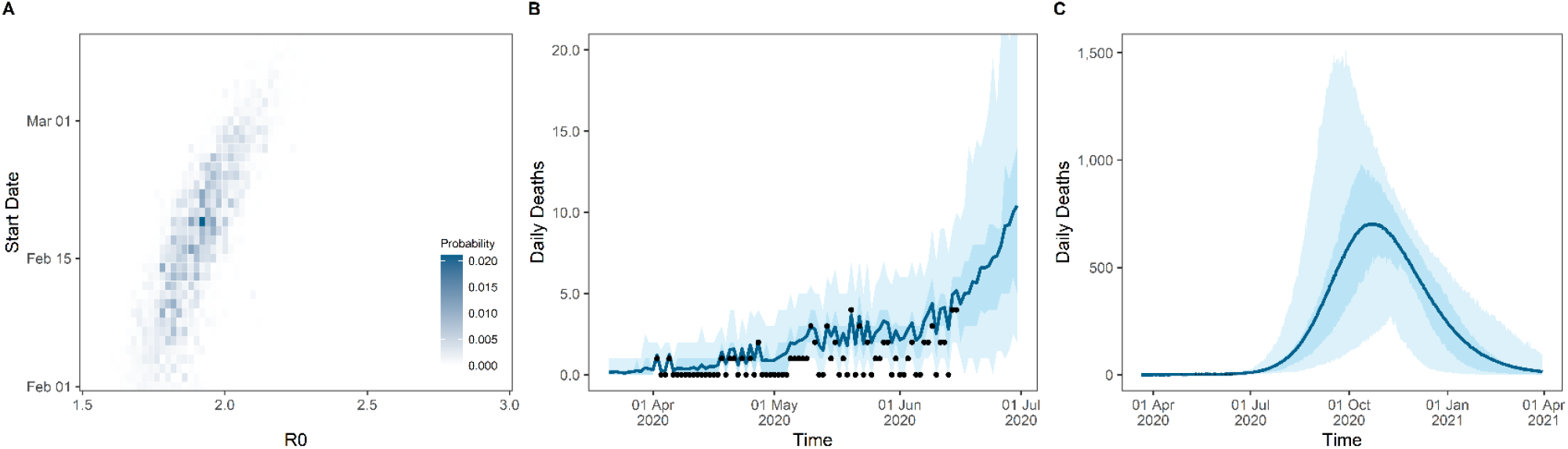
Model fitting to deaths time series assuming 50% reporting fraction. **A)** Grid search probabilities for each pair of potential epidemic start dates and R_0_ prior to intervention introduction. **B)** Model fit to daily death incidence. Black dots represent the reported number of deaths **C)** Model projections of deaths assuming no change in the current measures. Solid line represents the mean value of 100 model simulations, solid area colour represents the 50% range and shaded areas 95% range from the 100 model simulations.

Our estimates of the age distribution of all infections to date are similar to those reported (Figure 1C & Additional file 1: Figure S4A) suggesting that the age-based contact patterns driving the underlying attack rate in our model are appropriate for this setting. Based on our understanding of the severity of infection by age from China and Europe, we predict that severe cases requiring hospitalisation will disproportionately affect older age groups (Additional file 1: Figure S4B). This is consistent with the deaths that have occurred in older age-groups, with 77% of all deaths reported from those aged over 60 by 15th June.^20^

### Future Scenarios

Figure 3 shows the potential epidemic trajectories under the first four scenarios. Under scenario 1 we predict a significant epidemic which peaks in October. This is because we estimate that R_t_ currently remains above 1. Such a scenario is predicted to result in healthcare capacity being overwhelmed for much of the epidemic, resulting in significant morbidity with an estimated 77,400 deaths (95% CI 55,270-100,700) by the end of the epidemic (Figure 3, Table 2).

**Table 2.** Estimates of number of infections and deaths, and peak demand on hospital beds and ICU beds over the course of the epidemic under different intervention scenarios. Numbers in brackets represent the 95% range of model estimates. Assumed capacity for hospital beds with oxygen supply 407 and ICU beds with mechanical ventilators 15.

| Intervention Scenario | Total number infected | Total number deaths | Peak hospital bed demand | Peak ICU bed demand | Factor increase hospital bed with high pressure oxygen capacity to match peak demand | Factor increase ICU capacity by to match peak demand | Month in which epidemic peaks (Infections) |
| --- | --- | --- | --- | --- | --- | --- | --- |
| Non-Triggered Scenarios |  |  |  |  |  |  |  |
| Scenario 1 | 6776600<br>(4985100 - 8732200) | 77400<br>(55270 - 100700) | 4202<br>(1635 - 7956) | 344<br>(148 - 652) | 10 | 10 | October 2020 |
| Scenario 2 | 12217000<br>(11569000 - 12694000) | 139000<br>(132600 - 144200) | 15750<br>(7512 - 17520) | 1192<br>(583 - 1371) | 39 | 34 | October 2020 |
| Scenario 3 | 81166<br>(45028 - 164050) | 476<br>(248 - 1007) | 146<br>(78 - 278) | 35<br>(19 - 45) | 0 | 0 | June 2020 |
| Scenario 4 | 12511000<br>(11636000 - 13438000) | 142600<br>(133400 - 151900) | 16180<br>(3293 - 23430) | 1258<br>(266 - 1780) | 40 | 36 | January 2021 |
| Triggered Scenarios (following initial 3 month suppression period) |  |  |  |  |  |  |  |
| Scenario 5 | 260300<br>(182880 - 322820) | 716<br>(452 - 1036) | 172<br>(63 - 295) | 31<br>(15 - 57) | 0 | 0 | June 2020 |
| Scenario 6 | 308630<br>(45692 - 2322500) | 2242<br>(258 - 19800) | 150<br>(79 - 683) | 33<br>(20 - 71) | 0 | 0 | June 2020 |
| Scenario 7 | 6731600<br>(4963800 - 8648900) | 76300<br>(54490 - 99040) | 2743<br>(407 - 7423) | 229<br>(46 - 617) | 7 | 7 | March 2021 |

**Figure 3.**
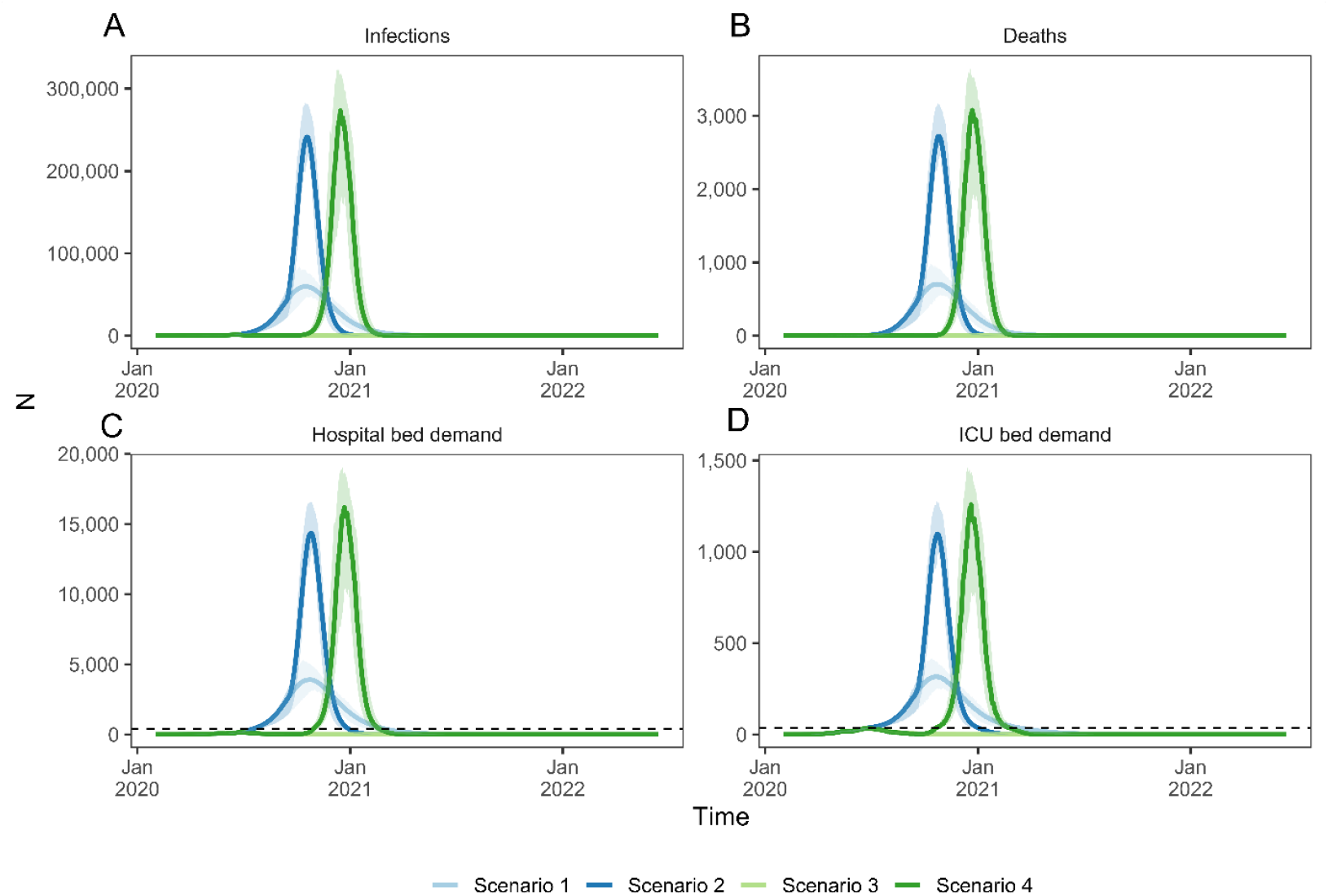
The impact of the four intervention scenarios compared with the current levels of mitigation. **A)** The daily incidence of new infections, **B)** the daily incidence of deaths, **C)** the daily number of hospital beds required and **D)** the daily number of ICU beds required. The current hospital bed capacity and ICU bed capacity are shown in black dashed lines in panels C and D respectively. The timing of peaks results from the different durations of scenarios we considered. Scenario 1 and scenario 3 are mitigation and suppression maintained long term and in scenario 2 and scenario 4 mitigation and suppression measures are relaxed after three months. Line is the mean value of 100 model simulations with shaded regions the 50% range.

If contact rates can be reduced by 60% compared to pre-epidemic levels (representing an additional 40% reduction on top of the average estimate from control measures to date) then the short term increases in cases would be much less rapid. If this level of suppression can be maintained (scenario 3), then we predict very few infections and deaths and healthcare demand would remain within capacity. The sustainability of such measures are crucially important – under the scenarios considered here, relaxation of reductions in contacts (scenarios 2 and 4) leads to rapid resurgence of the virus, resulting in substantial secondary peaks in the epidemic (Figure 3). In all scenarios apart from long-term suppression (scenario 3), the demand for hospital ad ICU beds is predicted to exceed current capacity which would need to increase up to 40 and 36 times respectively to meet the peak demand predicted in the worst-case scenario (Table 2).

Given these future scenarios and the current healthcare capacity we predict that substantial periods of time would need to be spent under suppression to relieve the demand on a fragile healthcare system. However, maintaining such an intense level of suppression is likely to have significant impacts on the local and national economy. We therefore additionally considered scenarios in which periods of suppression are triggered by healthcare demand or are less severe in their contact reductions.

Under these assumptions, we predict that periods of monthly suppression and a reduction in contacts outside of suppression could ease the pressure on the healthcare system and reduce deaths substantially (Figure 4, Table 2). In Scenario 5, given the current limited ICU capacity, suppression would need to be triggered at low incidence (six new cases requiring ICU care, sensitivity analysis to the trigger choice is shown in Additional file 1: Figure S5), and combined with enhanced suppression periods with a 75% reduction in contacts, to keep both general hospital and critical care services within capacity. Following these periods of enhanced month-long suppression, we estimate that it would take around 70 days to trigger the subsequent suppression period. This results in an additional 5-months of suppression, which will increase if post suppression contacts return to pre-epidemic levels (Additional file 1: Figure S6).

**Figure 4.**
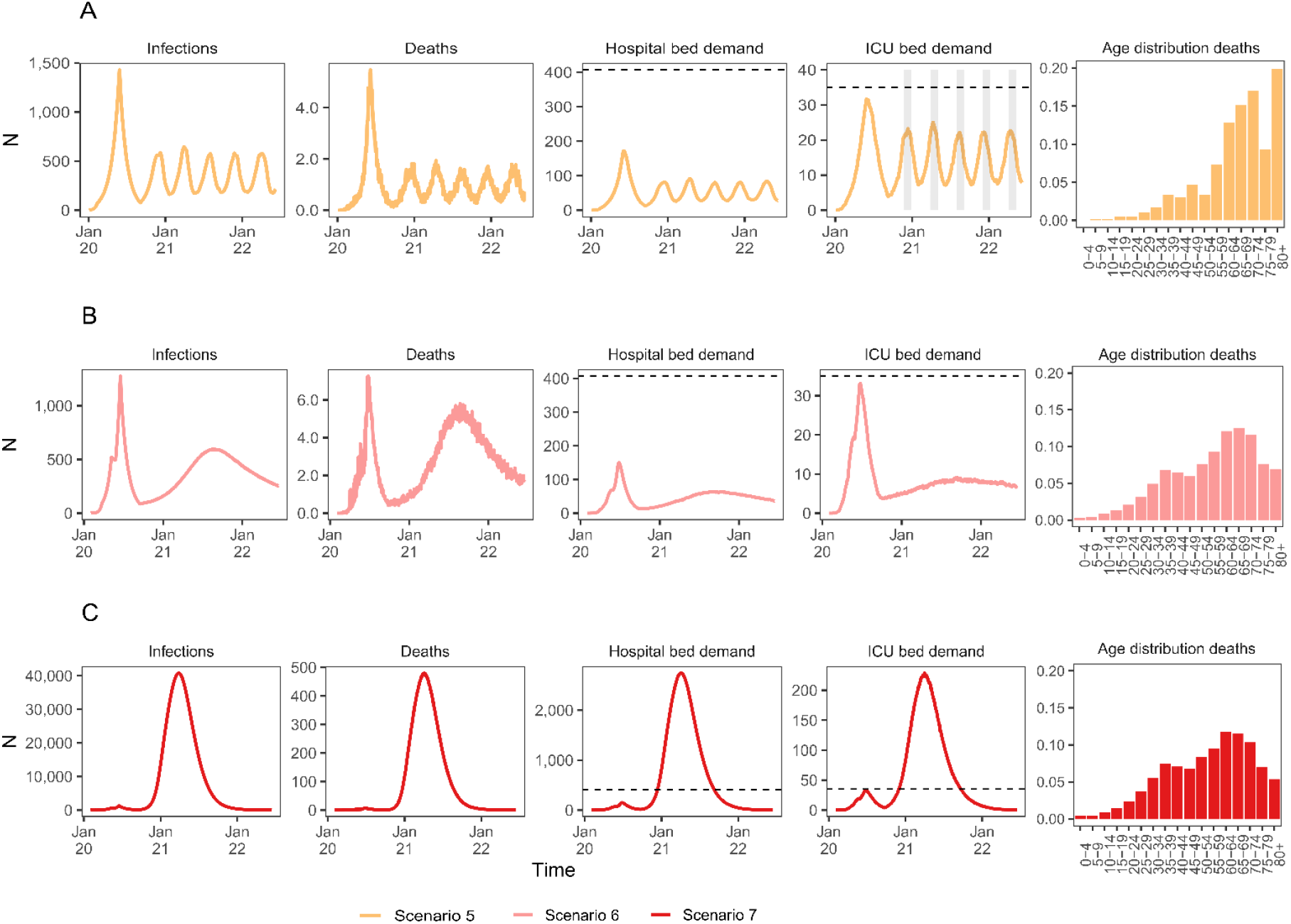
The impact of long term reductions in contacts on infections, deaths hospital bed and ICU bed demand and the age distribution of deaths. **A)** Scenario 5: months of triggered suppression at 75% reductions in contacts are represented by shaded grey bars. **B)** Scenario 6: reduction in contacts of 50% maintained long term after 3 months of suppression and **C)** Scenario 7: reduction in contacts by 33% maintained long term after 3 months of suppression. Dashed horizontal lines represent the current hospital and ICU bed capacity in Senegal. All interventions are assumed to remain in place for the duration of the simulation period. Line is the mean value of 100 model simulations.

Instead if contact reductions can be maintained at an intermediate level (scenario 6) we predict that demand for healthcare services remain below capacity (Figure 4). Given that R_t_ hovers around 0·95 in this scenario, this would protect the healthcare system and control transmission following the initial suppression period (Table 2). Finally, if following 3 months of suppression, contact patterns return to their current levels under the emergency measures (scenario 7), we predict a significant resurgence in cases and deaths (Table 2).

Age disaggregated proportions of hospitalised cases, and deaths are shown in Additional file 1: Figure S8 and Figure S9 respectively for all scenarios modelled. Whilst we predict that the burden of COVID-19 mortality will disproportionately affect the oldest age groups, we also note significant excess burden in those aged 30-60 (Additional file 1: Figure S7 & Figure S9). This is predicted to occur because of our assumptions regarding lower quality healthcare in this setting combined with an overall low capacity of hospital beds to match the required demand. For comparison Additional file 1: Figure S9B shows the distribution of deaths assuming the same quality care as in China, Europe and the US. Thus, scenarios that avoid capacity being overwhelmed are predicted to be particularly beneficial for the 30-60 age-group who remain at risk of exposure and for whom a significant subset are likely to require oxygen support.

## Discussion

Our results indicate that, through prompt action, Senegal has managed to significantly reduce transmission of SARS-CoV-2. Despite this, because we estimate the reproduction number remains above the threshold value of 1, there remains the potential for a severe epidemic that could overwhelm local health services and result in substantial excess mortality. A large driver of these mortality estimates is the explicit inclusion in the model of hospital capacity constraints to treat severe infections. In our model this leads to large excess deaths especially in younger age groups. Current trends in hospital bed occupancy from Senegal suggests that capacity is already close to being exceeded. For example, in Dakar on 2^nd^ June 98% of hospital beds were already occupied by COVID-19 patients.^21^ However, it is important to note that these scenarios are highly uncertain, and future trajectories will therefore very much depend on both the measures adopted and the impact that relaxing measures has on SARS-CoV-2 transmission, which will become clearer in the coming months.

Several commentaries have pointed to the reported slower growth of the COVID-19 epidemic across many African nations, which might be due to the early introduction of emergency measures or as yet uncharacterised differences.^8,22,23^ Given the trends observed to date we also predict slower growth with the epidemic peaking in October. However, due to the low levels of healthcare capacity, all but the most stringent scenarios considered predict that capacity to treat severe patients will be overwhelmed and incur significant excess mortality. In this context, using the time “bought” by suppressive measures to increase the availability of healthcare relevant to COVID-19 treatment, especially oxygen concentrators to treat those patients who do not require mechanical ventilation, will be invaluable to reduce poor patient outcomes. Senegal has already begun increasing its capacity to provide beds to mild patients and have repurposed hotels to allow for isolation of identified cases not requiring care.^24^ As patient numbers continue to rise in Senegal, rapidly addressing shortages to treat patients will be vital.

Without substantial surges in healthcare capacity, our results suggest that periods of triggered suppression in conjunction with extensive social-distancing outside of suppression periods may be required to minimise deaths from COVID-19. However, we note that triggered suppression will require a highly co-ordinated surveillance and hospital network and its feasibility will also be determined by levels of access to healthcare. As such reductions in transmission need not involve widescale restrictions, but rather could be achieved by locally triggered physical distancing as has been undertaken in much of Asia.^25^ Whilst these measures will help control COVID-19, the effects of long-term suppression will have economic and social implications for communities, with impacts likely to be concentrated in the poorest and most vulnerable groups.^26^ Disruption to health systems due to COVID-19 and the associated impact on other disease programmes such as malaria and TB is another important concern.^27,28^ The practicality of implementing even localised restrictions is therefore likely to be challenging and the ability for many populations to effectively social distance (e.g. working-from-home) and engage in infection prevention and control practices (e.g. handwashing) remains a major challenge in the context of overcrowding, poverty and stratification of manual labour towards the poorest.^26,29^

There are alternative strategies available however to mitigate these potential impacts. In this analysis we have focussed only on interventions relating to reducing population contact patterns– we do not consider the role of other interventions such as increased testing rates, widescale contact tracing and isolation activities. Such interventions also have a place in a comprehensive response plan to COVID-19 and could reduce the need for suppression/high levels of physical distancing if implemented at scale and in a proactive manner.^30-32^ At the current time many African nations, including Senegal, are scaling up their testing capacity.^3^ However, current levels are not yet on the scale of many of the countries pursuing aggressive testing as a control strategy.^33^ In addition, trials have begun on a rapid diagnostic test, which if the sensitivity and specificity are high enough could be important for countries in Africa with limited laboratory capacity.^34^

Several other modelling studies have come to similar conclusions in other LMIC settings.^5,6^ Of note, our estimates of the current R_t_ in Senegal and hence total infections are similar to those in a recent study modelling SARS-CoV2 transmission in several African countries which estimated R_0_=2·4 (95% CI 2·1-2·7) and Rt=1·5 (95% CI 1·0-2·0) using a method which compared efficacy of interventions against those in South Korea.^7^ Our projections of future burden are, however, substantially higher than a recently published study of likely COVID-19 burden in Africa, in which total future infections and deaths in Senegal are 3,937,580 (25% of the modelled population) and 2,225 respectively (representing an infection fatality ratio of 0·057%).^8^ Whilst there are many differences in methodological approach between the two studies, the key driver of their substantially lower estimates is the a priori assumption that African populations are less susceptible to both infection itself, and its downstream sequelae, based on both a “socio-ecological index” and “vulnerability index”. Although the elements of these indices (household size, urbanicity, slum populations, weather, connectedness, sanitation, co-morbidities) are all factors that are relevant to transmission, the indices were not validated against observed epidemic trajectories of SARS-CoV-2 transmission nor other similar respiratory infections. Whilst the spread of any respiratory pathogen is likely to be slower in rural areas compared to urban areas (as has been observed, for example, in the U.S), we are not aware previous examples of complete protection from respiratory pathogens on the basis of these factors other than in the most isolated communities and hence any such assumptions should be treated with caution.^35^

There are several limitations to the analyses presented here. Firstly, we rely on reported deaths to calibrate the model and estimate R_0_ and how it changes in response to control measures. Due to the low number of deaths, all scenarios presented here are highly uncertain and this needs to be borne in mind. In addition, the extent of under-reporting of deaths from COVID-19, and how it varies across the country, remains unclear. Here, we assume a 50% death ascertainment rate; whilst our results are sensitive to the extent of under-reporting assumed, they do not qualitatively change our conclusions surrounding healthcare capacity exceedance and the need for more stringent control measures. Given the small number of deaths to date, we only undertook this analysis at a national level; however it is clear from the geographical concentration of cases that there will likely be regional differences in the scale of individual outbreaks, as we have seen in other countries experiencing more advanced epidemics, and therefore further work will be need to understand regional trends and as such adapt policy to local needs. Additionally, we currently use Google mobility data to estimate the impact of emergency measures on contact patterns. While mobile phone coverage in Senegal is increasing annually, penetration throughout the population remains low. In 2018 only 34% of the population owned a smartphone with internet access compared to around 82% in the United Kingdom.^36^ Systematic biases in mobile ownership – either by location (urban vs rural) or by income strata could therefore lead to inferred contact patterns not being representative of the wider population. Finally, important to note is that our current estimates of the severity of COVID-19 rely on data from high income countries where patterns of comorbidities relevant to COVID-19 are likely to differ.^6,13,37^ Understanding how patterns of comorbidities will shape the age-profile of COVID-19 severity in lower-income countries will be crucial to generating more refined estimates of the burden these settings are likely to face.

Our study highlights how in the absence of efficacious pharmaceutical interventions, longer term physical distancing and suppressive measures may be needed to prevent a significant COVID-19 epidemic in Senegal.

## Conclusions

In this work we have accounted for the current sets of emergency measures in place in Senegal along with healthcare capacity constraints and age-specific patterns of COVID-19 severity to provide long-term projections of the potential epidemic dynamics. Importantly, we highlight that the current measures to date have successfully slowed the rate of transmission in Senegal and highlight the potential trajectories of the epidemic if the current policy is relaxed or enhanced. Our estimates of the impact of the COVID-19 epidemic in Senegal along with earlier studies highlight a consideration for the evaluation of continued and enhanced physical distancing measures and the scale up of the healthcare system to cope with increasing levels of COVID-19 patients. These estimates are therefore crucial to enable countries to make informed decisions with regards to their response policy. In addition this work highlights that we still require close and careful monitoring of the situation in Senegal and other Sub-Saharan African countries to be able to refine our estimates as more data is made available and, to crucially understand the impacts of complex disease burdens among other factors on the severity of COVID-19 in the region.

## Data Availability

The model structure used in this analysis is available in the form of an R package squire available from https://mrc-ide.github.io/squire/. Publicly available data was used to calibrate the model including key epidemiological data from the Ministere de la Sante et de l'Action Sociale of Senegal, whose dashboard can be found at: https://cartosantesen.maps.arcgis.com/apps/opsdashboard/index.html#/260c7842a77a48c191bf51c8b0a1d3f6 and Google COVID-19 community mobility reports https://www.google.com/covid19/mobility/.

https://cartosantesen.maps.arcgis.com/apps/opsdashboard/index.html#/260c7842a77a48c191bf51c8b0a1d3f6

https://www.google.com/covid19/mobility/

## Availability of data and materials

The model structure used in this analysis is available in the form of an R package *squire* available from https://mrc-ide.github.io/squire/. Publicly available data was used to calibrate the model including key epidemiological data from the Ministère de la Santé et de l’Action Sociale of Senegal, whose dashboard can be found at: https://cartosantesen.maps.arcgis.com/apps/opsdashboard/index.html#/260c7842a77a48c191bf51c8b0a1d3f6 and Google COVID-19 community mobility reports https://www.google.com/covid19/mobility/.

## Competing interests

The authors declare that they have no competing interests

## Funding

HAT and CW acknowledge support from an MRC DTP funding grant. OJW, PGTW and ACG acknowledge support from The Wellcome Trust and UK Department for International Development. PGTW and ACG acknowledge Centre support from the UK Medical Research Council. SN would like to acknowledge support from the MRC Centre for Global Infectious Disease Analysis (MR/R015600/1) and the Imperial BRC Centre. The funders had no role in the data collection, data analysis, data interpretation, or writing of the report.

## Author Contributions

HAT, AM, BC, SN, ACG and SM conceived the study. AM, BC, SM and HAT collected and analysed epidemiological data. OJW, CW, PGTW, ACG designed and developed model methodology and the associated R package used in this analysis. SM, AM, BC, SN provided guidance and data on the current situation of the epidemic in Senegal, the response policy and contextual information on the health system. HAT performed analysis and wrote the first draft of the manuscript and all authors contributed to the redrafting of the manuscript. The Imperial College COVID-19 Response Team were involved in developing the underpinnings of the COVID models, obtaining and analysing data used in its construction and parameterisation.

## Acknowledgements

Imperial College COVID-19 response team.

## Notes

### Competing Interest Statement

The authors have declared no competing interest.

## References

1. World Health Organization. Coronavirus disease (COVID-19) Situation Report - 147. 2020.

2. Houreld KL, David; McNeill Ryan; Samuel, Granados. Virus exponses gaping holes in Africa’s health systems. 2020. https://graphics.reuters.com/HEALTH-CORONAVIRUS/AFRICA/yzdpxoqbdvx/ (accessed May 2020).

3. Ministère de la Santé et de l’Action Sociale. Situation du Covid-19 au Senegal 2020. https://cartosantesen.maps.arcgis.com/apps/opsdashboard/index.html#/260c7842a77a48c191bf51c8b0a1d3f6 (accessed June 2020).

4. Government Republique Du Senegal. Decree n ° 2020-830 of 23 March 2020 proclaiming a state of emergency on the national territory. 2020. https://www.sec.gouv.sn/lois-et-reglements/lois-et-d%C3%A9crets (accessed May 2020).

5. van Zandvoort K, Jarvis CI, Pearson C, et al. Response strategies for COVID-19 epidemics in African settings: a mathematical modelling study. MedRxiv 2020.

6. Walker PG, Whittaker C, Watson OJ, et al. The impact of COVID-19 and strategies for mitigation and suppression in low-and middle-income countries. Science 2020.

7. Diop BZ, Ngom M, Pougué Biyong C, Pougué Biyong JN. The relatively young and rural population may limit the spread and severity of COVID-19 in Africa: a modelling study. BMJ Global Health 2020; 5(5): e002699.

8. Cabore JW, Karamagi H, Kipruto H, et al. The potential effects of widespread community transmission of SARS-CoV-2 infection in the WHO African Region: a predictive model. BMJ global health 2020.

9. Wallinga J, Lipsitch M. How generation intervals shape the relationship between growth rates and reproductive numbers. Proceedings of the Royal Society B: Biological Sciences 2007; 274(1609): 599–604.

10. Cori A, Ferguson NM, Fraser C, Cauchemez S. A new framework and software to estimate time-varying reproduction numbers during epidemics. American journal of epidemiology 2013; 178(9): 1505–12.

11. Jombart T, Cori A, Nouvellet P. earlyR. https://www.repidemicsconsortium.org/earlyR/.

12. Melegaro A, Del Fava E, Poletti P, et al. Social contact structures and time use patterns in the Manicaland Province of Zimbabwe. PloS one 2017; 12(1).

13. Verity R, Okell LC, Dorigatti I, et al. Estimates of the severity of coronavirus disease 2019: a model-based analysis. The Lancet Infectious Diseases 2020.

14. Ministère de la Santé et de l’Action Sociale. Capacité litière des EPS cible pour la PEC des cas de covid-19 du 25 avril 2020. 2020.

15. Google. COVID-19 community mobility reports 2020. https://www.google.com/covid19/mobility/ (accessed May 2020).

16. Nouvellet P, Bhatia S, Cori A, et al. Report 26: Reduction in mobility and COVID-19 transmission. Imperial College London 2020.

17. Ainslie KE, Walters CE, Fu H, et al. Evidence of initial success for China exiting COVID-19 social distancing policy after achieving containment. Wellcome Open Research 2020; 5.

18. R Core Team. R: A language and environment for statistical computing. R Foundation for Statistical Computing. Vienna, Austria 2020.

19. Bi Q, Wu Y, Mei S, et al. Epidemiology and transmission of COVID-19 in 391 cases and 1286 of their close contacts in Shenzhen, China: a retrospective cohort study. The Lancet Infectious Diseases 2020.

20. Ministère de la Santé et de l’Action Sociale. Riposte à l’épidémie du nouveau coronavirus COVID-19, Sénégal. Rapport de situation n°27 du 8 juin 2020. 2020.

21. Centre des Opérations d’Urgence Sanitaire. Réunion de coordination de la Réponse à l’Epidémie de COVID-19 Ministère de la Santé et de l’Action Sociale; 2020.

22. Why Africa’s coronavirus outbreak appears slower than anticipated MedicalXpress 2020.

23. Anna C. Africa virus cases surpass 100,000; lockdowns slowed growth. abcNews 2020.

24. Shryock R. Senegal pledges a bed for every coronavirus patient — and their contacts, too. NPR. 2020.

25. Dighe A, Cattarino L, Cuomo-Dannenburg G, et al. Report 25: Response to COVID-19 in South Korea and implications for lifting stringent interventions. Imperial College London 2020.

26. Peter Winskill, Charlie Whittaker, Patrick Walker, et al. Report 22: Equity in response to the COVID-19 pandemic: an assessment of the direct and indirect impacts on disadvantaged and vulnerable populations in low- and lower middle-income countries. Imperial College London 2020.

27. Sherrard-Smith E, Hogan AB, Hamlet A, et al. Report 18: The potential public health impact of COVID-19 on malaria in Africa. Imperial College London 2020.

28. Hogan AB, Jewell B, Sherrard-Smith E. Report 19: The Potential Impact of the COVID-19 Epidemic on HIV, TB and Malaria in Low-and Middle-Income Countries. Imperial College London 2020.

29. Loayza N. Smart containment and mitigation measures to confront the COVID-19 pandemic: Tailoring the pandemic response to the realities of developing countries 2020. https://blogs.worldbank.org/developmenttalk/smart-containment-and-mitigation-measures-confront-covid-19-pandemic-tailoring (accessed May 2020).

30. Kucharski AJ, Klepac P, Conlan A, et al. Effectiveness of isolation, testing, contact tracing and physical distancing on reducing transmission of SARS-CoV-2 in different settings. medRxiv 2020.

31. Grassly N, Pons Salort M, Parker E, et al. Report 16: Role of testing in COVID-19 control. 2020.

32. Hellewell J, Abbott S, Gimma A, et al. Feasibility of controlling COVID-19 outbreaks by isolation of cases and contacts. The Lancet Global Health 2020.

33. Roser M, Ritchie H, Ortiz-Ospina E, Hasell J. Coronavirus Pandemic (COVID-19). 2020. https://ourworldindata.org/coronavirus (accessed May 2020).

34. Haque N. Senegal trials begin for $1 COVID-19 test kit. Aljazeera. 2020.

35. Unwin H, Mishra S, Bradley V, et al. Report 23: State-level tracking of COVID-19 in the United States. Imperial College London 2020.

36. Silver L, Johnson C. Internet Connectivity Seen as Having Positive Impact on Life in Sub-Saharan Africa: But Digital Divides Persist: Pew Research Center; 2018.

37. Clark A, Jit M, Warren-Gash C, et al. How many are at increased risk of severe COVID-19 disease? Rapid global, regional and national estimates for 2020. medRxiv 2020.

38. Ministère de la Santé et l’Action sociale. Riposte à l’épidémie du nouveau coronavirus COVID- 19, Sénégal. Rapport de situation n°26 du 04 juin 2020. 2020.

